# Genomic Research as a Critical Pathway to Diagnosis for Individuals with Rare Disease

**DOI:** 10.64898/2026.09.15.26363084

**Authors:** Anne O’Donnell-Luria, Stephanie DiTroia, Melanie C. O’Leary, Lynn Pais, Vijay S. Ganesh, Gabrielle Lemire, Emily O’Heir, Ashana Neale, Alicia Pham, Grace E. VanNoy, Brian E. Mangilog, Moriel Singer-Berk, Alba Sanchis-Juan, Hana Snow, Siwaar Abouhala, Stacey Agu, Mutaz Amin, Samantha M. Baxter, Benjamin Blankenmeister, Kinga M. Bujakowska, Colleen M. Carlston, Sanaa Choufani, Laura E. Covill, Eleina M. England, Carmen Glaze, Julia Goodrich, Emily Groopman, Kristen Laricchia, Alysia Kern Lovgren, Jialan Ma, Eva Martínez, Chloe Mighton, Briana O’Leary, Ikeoluwa A. Osei-Owusu, Sander Pajusalu, Lindsay Romo, Kathryn Russell, Riccardo Sangermano, Eleanor Seaby, Jillian Serrano, Gulalai Shah, Mugdha Singh, Kayla M. Socarras, Sarah L. Stenton, Miriam S. Udler, Ben Weisburd, Rosanna Weksberg, Clara E. Williamson, Rare Genomes Project, Niall Lennon, Christina Austin-Tse, Harrison Brand, Michael E. Talkowski, Daniel G. MacArthur, Monica H. Wojcik, Heidi L. Rehm

**Affiliations:** Program in Medical and Population Genetics, Broad Institute of MIT and Harvard, Cambridge, MA, 02142, USA; Division of Genetics and Genomics, Boston Children’s Hospital and Harvard Medical School, Boston, MA, 02115, USA; Center for Genomic Medicine, Massachusetts General Hospital, Boston, MA, 02114, USA; Harvard Medical School, Boston, MA, 02115, USA; Department of Neurology, Mass General Brigham, Boston, MA, USA; Ocular Genomics Institute, Mass Eye and Ear and Harvard Medical School, Boston, MA, 02114, USA; Genetics and Genome Biology, Hospital for Sick Children Research Institute, Toronto, ON, M5G 1X8, Canada; Manton Center for Orphan Disease Research, Boston Children’s Hospital, Boston, MA, 02115, USA; Division of Maternal Fetal Medicine, Department of Obstetrics and Gynecology, Brigham and Women’s Hospital, Boston, MA, 02115, USA; Genetics and Personalized Medicine Clinic, University of Tartu and Tartu University Hospital, Tartu, 50406, Estonia; Department of Genetics, Yale School of Medicine, New Haven, CT, 06520, USA; Endocrine Division, Department of Medicine, Massachusetts General Hospital, Boston, MA, 02114, USA; Broad Institute of MIT and Harvard, Cambridge, MA, 02142, USA; Broad Clinical Labs, Broad Institute of MIT and Harvard, Burlington, MA, 01801, USA; Division of Newborn Medicine, Boston Children’s Hospital, Boston, MA, 02115, USA; Department of Pathology, Harvard Medical School, Boston, MA, 02115, USA

## Abstract

Many individuals with rare monogenic disease remain molecularly undiagnosed due to challenges accessing genetic testing, ambiguity in interpretation of uncertain variants, and latency between novel disease-gene discovery and adoption into clinical pipelines. The Rare Genomes Project (RGP) provides a remote, research-based genomic sequencing (GS) service for rare disease families to overcome diagnostic barriers while advancing the understanding of the genetic basis of rare disease. Short-read GS and family-centered analysis were performed to identify causes of primary disease. Variants meeting return criteria were clinically confirmed and returned to families. Since 2017, RGP enrolled clinically heterogeneous, undiagnosed rare disease families from all fifty states, Washington DC, and Puerto Rico. Findings were returned to 272 of 1014 sequenced families, resulting in a solve rate of 23.3% and a returned strong candidate rate of 3.5%. Condition-specific return rates ranged from 15% to 40% with many families receiving a result in a known disease gene after healthcare access challenges. Research interventions contributed to returns in 16% of all families and included variants in recently discovered or non-coding disease genes, additional functional studies to resolve variant uncertainty, and real-time data sharing to empower novel disease-gene discovery. RGP demonstrates that a research-based, patient-driven remote sequencing study can effectively bridge a diagnostic gap for rare disease families underserved in current clinical care. This study also underscores the clinical utility of GS with frequent re-analysis and highlights the critical continued role of research in variant interpretation, method innovation, and genomic knowledge base expansion for rare disease diagnosis.

## INTRODUCTION

Despite rapid advancements in genomic technologies and increasing adoption of genome sequencing in clinical care, the majority of rare disease patients still lack a genetic diagnosis^1–3^. A diagnosis has many medical and social implications^4,5^, but barriers to testing and diagnosis continue to exist. Barriers include a combination of restrictive insurance coverage, difficulty accessing specialized genetics care, an overwhelming range of genetic testing options, delays in the adoption of the latest scientific knowledge and technologies for clinical diagnostic laboratories, and insufficient evidence to resolve variants of uncertain significance^6–9^. Additionally, the majority of the estimated 6,000-14,000 monogenic gene-disease relationships that are yet to be identified^10^ will require research investigation, collaboration, and detailed individual-level data sharing, often difficult to achieve in the clinical care setting.

Multiple US-based research programs exist to help undiagnosed rare disease families, including the NIH Undiagnosed Diseases Program (UDP) started in 2008 and later expanded into the Undiagnosed Disease Network, and the NHGRI-funded Centers for Mendelian Genomics in 2011 that was succeeded by the Genomics Research to Elucidate the Genetics of Rare diseases (GREGoR) consortium. These two major NIH-funded research programs have demonstrated that comprehensive clinical and genomic evaluation provide diagnoses to individual families and also empower new disease-gene discoveries, often possible only through international collaborative efforts in the investigation of rare disorders^11–13^. Likewise, similar programs in other countries span geographical regions around the world^14–18^. While these studies have contributed significant advancement to the clinical diagnosis of rare disease patients, they are often geographically limited and/or leave gaps in access for families unable to fulfill the requirements of an in-person assessment or physician referral^19^.

The Rare Genomes Project (RGP; raregenomes.org) was launched in 2017 by the Broad Institute of MIT and Harvard as a research program aimed at removing barriers in obtaining diagnoses for rare disease families with suspected monogenic conditions across the United States. The fully remote, direct-to-patient study design leveraged partnerships with families and advocacy groups to provide access to genomic sequencing, emerging technologies (e.g., long-read sequencing, RNA-seq, AI-based analysis), ongoing re-analysis, and rare variant classification and interpretation without the requirements of clinician referral or in-person visits. RGP treats genomic data as a sustained research asset, rather than a snapshot test analyzed once, with periodic integration and reanalysis of genomic data with longitudinal clinical data.

In this paper, we describe RGP’s experience recruiting a heterogeneous cohort of individuals across the U.S. actively seeking a genetic diagnosis for their rare disease, and summarize demographics, applications, and enrollment numbers. We report the diagnostic utility of short-read genomic sequencing (GS) and supporting technologies, highlighting areas where obtaining a genetic diagnosis could simply be addressed through improved access to currently available clinical testing. This work explores the ongoing critical need for rare disease genomic research programs to drive disease-gene discovery, to innovate new clinical sequencing techniques and analytic methodologies, and to provide ethically consented datasets to accelerate the advancement of rare disease discovery and diagnosis.

## SUBJECTS AND METHODS

### Recruitment, eligibility, and application

The study was approved through the Mass General Brigham IRB (Protocol# 2016P001422). RGP utilized a wide variety of recruitment methods including social media, provider referrals, and partnerships with rare disease advocacy organizations, including multiple groups of the Rare As One network (https://chanzuckerberg.com/science/programs-resources/rare-as-one/network/). Eligibility criteria for the program included having a genetically undiagnosed suspected monogenic disease (excluding hereditary cancer), access to a telephone and/or computer, a local medical provider to assist with clinical confirmation testing, and residency in the United States, its territories, or the federal capital district. Eligibility was initially limited to English-speaking individuals, was later expanded to include Spanish language speakers in year four, and then again expanded to include all languages in year five.

Prospective participants or a family member/guardian completed an application online or by phone (Appendix 1). Requested information included a brief narrative summary of the medical condition, age of symptom onset, prior medical work-up and genetic testing, and the affected status and availability of first-degree relatives. Demographic information, including state/district/territory of residence and self-reported race and ethnicity, was also collected. Alternatively, some medical providers obtained permission to send information on behalf of individuals through a referral form and to have study staff initiate contact about completing an application. Applicants were classified as “provider-referred” if they indicated that they heard about RGP from a medical provider or if a medical provider sent referral paperwork; all other applications were classified as “self-referred”.

To evaluate the probability of a monogenic disease etiology, all applications were subjected to a dual review by medical geneticists and/or genetic counselors. After dual review, eligible families were then invited to enroll. In cases where applications lacked sufficient medical details, probands were invited to enroll for a medical record review before determination of eligibility.

### Enrollment and consent

Enrollment was offered to the proband, the biological parents, affected extended family members, and up to two unaffected siblings. Video or telephone informed consent sessions were conducted with each individual and included consent to sample collection, medical record collection, recontact, return of primary findings or incidental findings, and the sharing of de-identified phenotype, biospecimen, and genomic data. The initial data sharing consent level was health/medical/biomedical use (HMB) at study outset and later expanded to general research use (GRU) in year four.

### Sample and medical record collection and processing

Human Phenotype Ontology (HPO) terms were extracted from the health history narrative provided in the application. The medical records of affected individuals were requested for review during genomic analysis. Self-reported prior testing was categorized into four groups: “genomic” (exome sequencing (ES) and/or genome sequencing (GS)), “some” (any other genetic test(s) exclusive of ES/GS), “none” (no prior genetic testing), or “unknown”. Families were deemed to have faced clinical testing access issues if they self-reported no prior testing, explicitly stated they could not obtain clinical testing, were provider-referred without prior ES or GS, or had medical record documentation of provider intent to order additional genetic testing that was not performed.

Blood sample collection kits containing an EDTA tube, paperwork for phlebotomy services, and return shipping materials were mailed directly to each participant. Kits also included a PAXgene RNA tube in year 5 onwards. Blood draws were performed at a local facility of the family’s choosing. In a small subset of cases where blood collection posed a challenge, a mobile phlebotomy service was used or alternative sample types were collected including saliva, buccal, or previously extracted DNA. When available, fresh-frozen, disease-relevant tissue from medically-indicated clinical procedures was also collected from affected participants.

Blood, saliva, and buccal samples underwent non-CLIA DNA extraction and quantification at Broad Clinical Labs or an RGP clinical lab partner. Research short-read GS was performed on an aliquot of extracted DNA from the proband and, if available, the biological parents and additional affected family members with residual DNA stored for future studies. DNA from enrolled unaffected family members was stored for possible future segregation studies. PAXgene RNA tubes were frozen for possible future extraction and sequencing.

### Short-read genome and transcriptome sequencing

RGP short-read genome and transcriptome sequencing was performed at Broad Clinical Labs. Genome sequencing libraries were created using PCR-free preparation of sample DNA (350 ng input at >2 ng/ul) and Illumina HiSeq X Ten v2 chemistry. Libraries were sequenced to a mean target coverage of >30x. Genomic data was originally processed with GATK workflows, but since, all data has been either reprocessed or initially processed with DRAGEN v3.7.8 for SNV (Single Nucleotide Variant) and indel (insertion/deletion) calls. Genomic VCF (Variant Call Format) files were joint-called using Broad’s Genomic Variant Store (GVS) workflow.

Additional calling and annotation pipelines were run on genomes to maximize diagnostic breadth and yield. These pipelines include GATK-SV and GATK SVAnnotate to detect and annotate structural variants (SVs)^20^; MitoSAlt and the gnomAD-mitochondrial DNA variant calling pipeline to identify large deletions and SNV/indels in the mitochondrial DNA (mtDNA)^21^; ExpansionHunter v5 to identify known disease-associated tandem repeat expansions (TREs)^22^; and SMA Finder to identify potential cases of spinal muscular atrophy due to deletions or gene conversions resulting in *SMN1* deficiency^23^.

For a subset of affected individuals, primarily those with variants suspected to impact splicing and those undergoing lrWGS, transcriptome sequencing (RNA-seq) was also performed on blood or disease-relevant frozen tissue samples. RNA-seq was completed using poly(A)-selection of mRNA transcripts and strand-specific cDNA library preparation. Libraries with a mean insert size of 550 bp were sequenced on the HiSeq 2500 platform to a minimum depth of 75-150 million STAR-aligned reads. External RNA Controls Consortium (ERCC) RNA spike-ins were used to control for sample variability.

### Additional data types

Additional genomic sequencing was performed on a subset of individuals based on sample availability, phenotype, and absence of causal variants identified in short-read genome data. The data generated depended on the specific collaboration and techniques being performed. Many of the methods and techniques applied were published previously, including: research-grade DNA methylation arrays^24^, long-read Oxford Nanopore or PacBio DNA sequencing^25,26^, long-read RNA isoform sequencing (ONT or MAS-ISO-seq/Kinnex on PacBio)^26^, and optical genome mapping^27^. Participants with additional data types are reported in Table S2, with technology combinations shown in Figure S1.

### Case analysis and matchmaking

Individual-level data, including phenotypes and genomic and transcriptomic data, were loaded into the *seqr* platform for family-centered analysis of SNVs, indels, and SVs^28^ and complemented by TRE and SMAFinder analysis external to *seqr*. Analyzed family structures were categorized as proband+ (proband with or without non-parent relatives), duo+ (parent-child duos with or without non-parental relatives), and trio+ (parent-child trios with or without additional relatives). Using *seqr*, a genomic analyst performed four preset family-based searches outlined in Table S3 and prioritized variants based on the *seqr* displayed annotations from *in silico* predictors (e.g., CADD, REVEL, and SpliceAI) and numerous rare disease online resources (e.g. GenCC, PubMed, ClinVar, Human Gene Mutation Database (HGMD), MONARCH) as described previously^28^. If candidate or diagnostic variants were not identified in the preset restrictive and permissive searches, search criteria were expanded further to assess variants outside of traditional Mendelian disease inheritance modes (e.g., incomplete penetrance, parental mosaicism) to maximize yield.

Candidate variants in uncharacterized disease-genes were submitted to Matchmaker Exchange (MME), a decentralized network of genomic matchmaking nodes built to support the sharing of data needed for novel disease-gene discovery^29^, via the *seqr* node. Matches were continually assessed for genotype and phenotype overlap and successful matches pursued for evidence aggregation and publication with participant consent.

### Defining GS added yield

To calculate the added yield of GS over ES, variants were determined to require GS if they were in non-coding genes, more than 20 bases from the intron/exon boundary, deletions or duplications of fewer than 3 exons, complex structural variants, or TREs, to allow direct comparison to previous work^30^. All other variants were considered detectable by ES, including *SMN1/SMN2* [MIM:600354/601627], copy number variants, and mitochondrial variants. Unlike Wojcik et al 2024, exome data for RGP participants was not available to assess individual sample limitations.

### Variant and gene curation

Candidate and diagnostic variants were classified according to ACMG/AMP standards with additional guidance provided by the Clinical Genome Resource (ClinGen) (https://clinicalgenome.org/working-groups/sequence-variant-interpretation/^31,32^. In addition to variant interpretation, undercharacterized gene-disease associations were classified according to ClinGen’s gene-disease validity framework^33^. Completed curations were submitted to ClinVar (for variant classifications) or GenCC (for gene classifications) by the Broad Center for Mendelian Genomics (Broad CMG; ClinVar Organization ID: 506627; GENCC:000115).

### Diagnostic yield and solve status

Diagnostic yield was determined by combining assessments of gene-disease validity, variant pathogenicity, inheritance, and phenotypic overlap between the proband and disease phenotypes or representative cases in the literature, as previously defined by the GREGoR consortium data model as Solve Status^34^. A case was considered “solved” or “probably solved” when pathogenic, likely pathogenic, or high evidence variants of uncertain significance (VUS-high: variants with one piece of supporting or moderate evidence away from likely pathogenic) were found in a known or novel disease gene with moderate or higher level of gene-disease validity and a clear clinical overlap with the RGP participant’s phenotype. A third category, “unsolved with a candidate”, included cases with insufficient support for variant pathogenicity and/or gene-disease validity and cases in which a pathogenic variant was identified in a gene that did not clearly fit participant’s phenotype or inheritance pattern, representing possible phenotype expansions. The final category, “unsolved”, defined cases with no clear candidate after analysis. Iterative manual re-analysis of “unsolved” and “unsolved with a candidate” cases remains ongoing, as well as continual automated re-analysis of all RGP cases in *seqr* using methods developed in concert with Talos^35^.

A small number of cases had a solve status adjustment after returning the result to a participant, and those adjustments were determined by a consensus among the multidisciplinary team and the participant’s clinical provider. Examples of solve status changes included cases where the physician did not feel the diagnosis was a phenotype match (resulting in reduced evidence adjustment) or provided additional clinical details that supported the diagnosis (resulting in higher evidence adjustment). Solve status reported here is up to date as of manuscript submission.

### Returnable criteria and return of results

Candidate and diagnostic variants identified as plausibly causal for an RGP participant’s phenotype were presented at a weekly multi-disciplinary case conference of clinicians, clinical laboratory geneticists, genetic counselors, genomic analysts, research trainees, and computational scientists. Variants meeting “solved” or “probably solved” criteria were automatically deemed returnable. Candidate variants were flagged for return on a case-by-case basis. Reasons for returning a candidate variant included unresolvable variants of uncertain significance (VUS) in a gene/condition for which other clinical tests are diagnostic (e.g., plasma amino acids), VUS-high variants with good clinical correlation, and candidates to be included in a novel gene discovery publication. Significance testing for categorical variables associated with receiving a returnable result was performed using the built-in logistic regression model in R version 4.4.1 (2024-06-14). Statistical significance was defined as p < 0.05

For candidate variants that did not meet consensus for return, next steps were discussed including: collection of additional medical information, sequencing additional family members, performing additional -omics testing (e.g., RNA-seq on blood, a derived cell line or other clinically available tissue), and identifying collaborations for additional variant-to-function modeling options.

Families were first notified of a returnable result and given the option to proceed with clinical confirmation. Clinical confirmation was performed through a variety of methods, including Sanger sequencing, ddPCR, gene-specific GS, gene-specific TRE analysis, mitochondrial DNA sequencing, chromosomal microarray, and *SMN1/SMN2* dosage analysis, prior to disclosure to participants. For one result where no gene-specific clinical test existed (*NOTCH2NLC* [MIM:603472]), non-CLIA research confirmation was performed by a collaborator through an orthogonal method of repeat-primed PCR (RP-PCR) and fluorescence amplicon length analysis (AL-PCR). Confirmation testing for all returns was coordinated by a study genetic counselor and included samples from the proband, as well as other family members when needed to confirm phase or *de novo* status. Testing was officially ordered by a local medical provider of the family’s choosing or, less commonly, a third-party telemedicine genetics provider, who disclosed the results to the family. After disclosure, a follow-up call was conducted with the family to address remaining questions about the study findings and to ensure the families were made aware of relevant support organizations, websites, and resources. Families were referred back to their clinician or a telemedicine provider to address questions regarding medical care.

Pathogenic variants in ACMG SF v3.2 genes^36^ not previously known to the family that were identified incidentally during analysis for the primary phenotype were returned clinically through the same return of results process after IRB consultation, per our protocol.

## RESULTS

### Applications and Enrollment

RGP received 2390 complete applications from May 2017 through January 2026. More than half of the families self-initiated an application to RGP (57%) predominantly completed by probands (n=785) or by parents/guardians of probands under age 18 or adult-aged probands unable to provide consent (n=575). An additional 37% of families self-applied to RGP after discussion with a medical provider. Applications from the remaining 5% of families were initiated by formal referral from the family’s provider. Nine percent of families who discussed RGP with a provider and 41% of families self-initiating an application did not meet eligibility criteria (28% overall), with the most common reasons being low suspicion for a monogenic disease, the need for clinical assessment to discern primary versus secondary phenotypes, and a known molecular diagnosis that explained the key phenotypes listed in the application. Eighteen percent of applications necessitated additional medical record review as the initial application did not contain sufficient information to assess the likelihood of a monogenic condition or to ascertain whether the prior genetic testing was non-diagnostic, after which 60% met eligibility criteria and were invited to enroll. In total, 1433 (60%) of all applications received were eligible to participate (Figure 1A).

**Figure 1:**
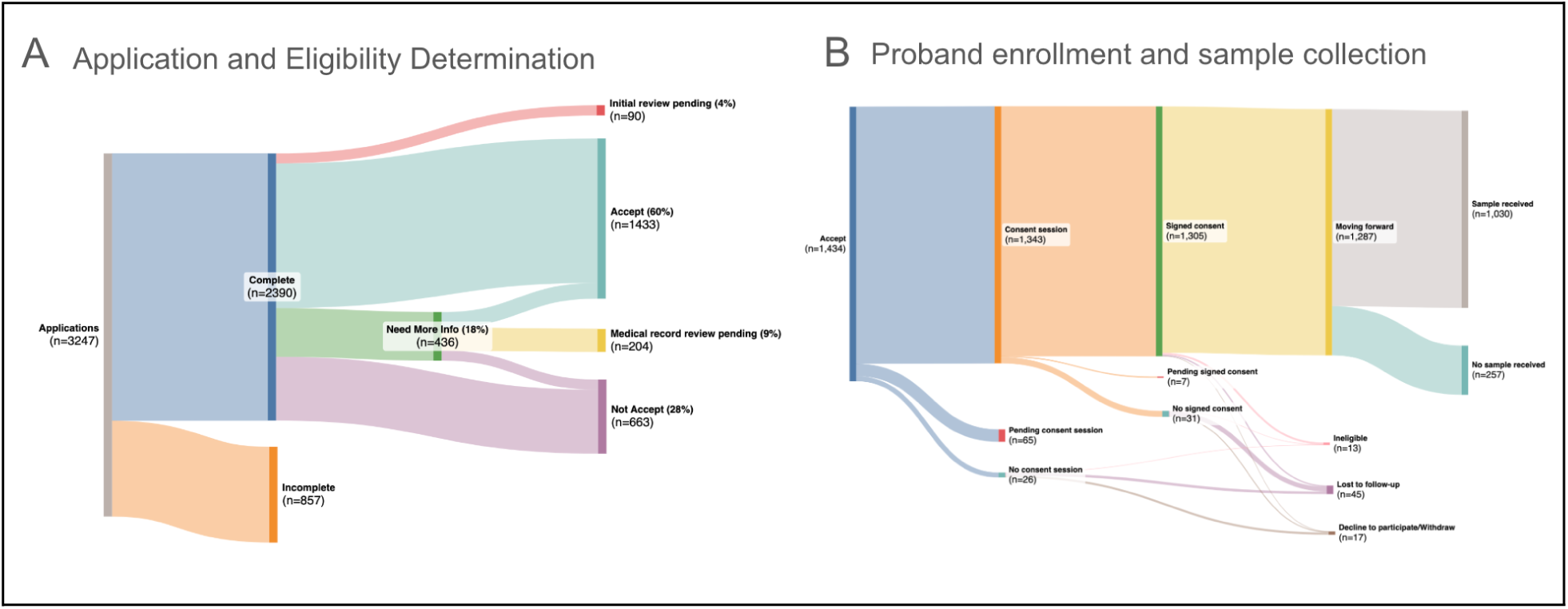
Applications, enrollment, and proband demographics. (A) **Application and Eligibility Determination** Sankey diagram of applications submitted and study eligibility determination of completed applications. Eligibility assessment decisions include Accept, Need More Info, and Not Accept. After medical record review, probands enrolled as Need More Info become Accept or Not Accept. (B) **Proband enrollment and sample collection** Sankey diagram of probands meeting eligibility criteria. The majority of accepted probands (72%) participated in a consent session, signed an informed consent document, and submitted a sample for sequencing. The remaining probands had not yet submitted samples, were lost to follow-up, deemed ineligible based on new information, or declined to participate/withdrew.

Ninety-one percent of accepted families participated in a video or telephone informed consent session, resulting in enrollment of 3478 individuals from 1305 families. Five percent of enrolled families (n=75) were lost to follow-up, withdrew from the study, or no longer met eligibility criteria. In total, 1287 families remained engaged with the study, and 80% of these probands (n=1030) provided a sample for sequencing (Figure 1B).

Nearly a third of accepted families (30%; 435/1433) faced barriers accessing clinical testing as indicated by one or more of the following: self-report of no prior testing for the suspected monogenic condition (n=126), explicit statements about an inability to obtain testing (n=40), referral by a clinical provider without prior ES or GS (n=297), or medical record documentation of intent to order additional testing that was not performed (n=68). This 30% may be an underestimate, however, as the presence or absence of access issues was not an explicit question posed to all applicants, but was instead inferred from information provided on the RGP application (Appendix 1) or through medical record review. An additional 303 of accepted families (21%) were referred by providers due to inability to obtain clinical GS after a negative or inconclusive clinical ES.

### Sequenced Cohort Demographics

RGP participants reside in all 50 states plus the District of Columbia and Puerto Rico, and demographic characteristics of probands who underwent GS are shown in Table 1. While proband age at enrollment was approximately split between adults and minors, the majority reported a childhood age of disease onset (72%). Many probands self-reported or had medical records documenting genetic testing prior to enrollment (88%), half of which reported prior ES/GS and half reported non-genomic testing (e.g., gene panel, microarray, or karyotype) (Table 1). Accuracy of self-reported prior genetic testing demonstrated high concordance rates when comparing application narrative to proband medical records across a subset of families; values were corrected when known to be discordant with medical records (Table S1).

Enrolled proband phenotypes spanned numerous organ systems and medical specialties, with the majority being complex neurodevelopmental (32%, n=330) or neuromuscular (30%, n=307) (Figure S2A). The age of onset varied significantly by phenotype, with seizure and neurodevelopmental presentations skewed toward pediatric onset, and neuromuscular disorders skewed toward adult onset (Figure S2B).

### Genomic Analysis

Iterative GS analysis on 1,014 families identified 575 variants of interest (Table S4). At time of publication, 346 variants in 272 families met study criteria for return (see METHODS), resulting in a cohort yield of 27.4% “solved”, “probably solved”, and returned VUS-high candidate cases (Figure 2A). Sixty-two percent of the variants returned reached pathogenic or likely pathogenic classification using ACMG/AMP guidelines^32^, and 83% were in genes with gene-disease relationships curated at moderate or above using ClinGen’s gene-disease validity framework^33^ (Figure S3, Table S2). Variants that did not meet return criteria were shared through Matchmaker Exchange, ClinVar, and the GREGoR genetic findings dataset with the intention of collecting additional evidence to support or refute causality. A full list of RGP prioritized and/or returned variants can be found in Table S4.

**Figure 2:**
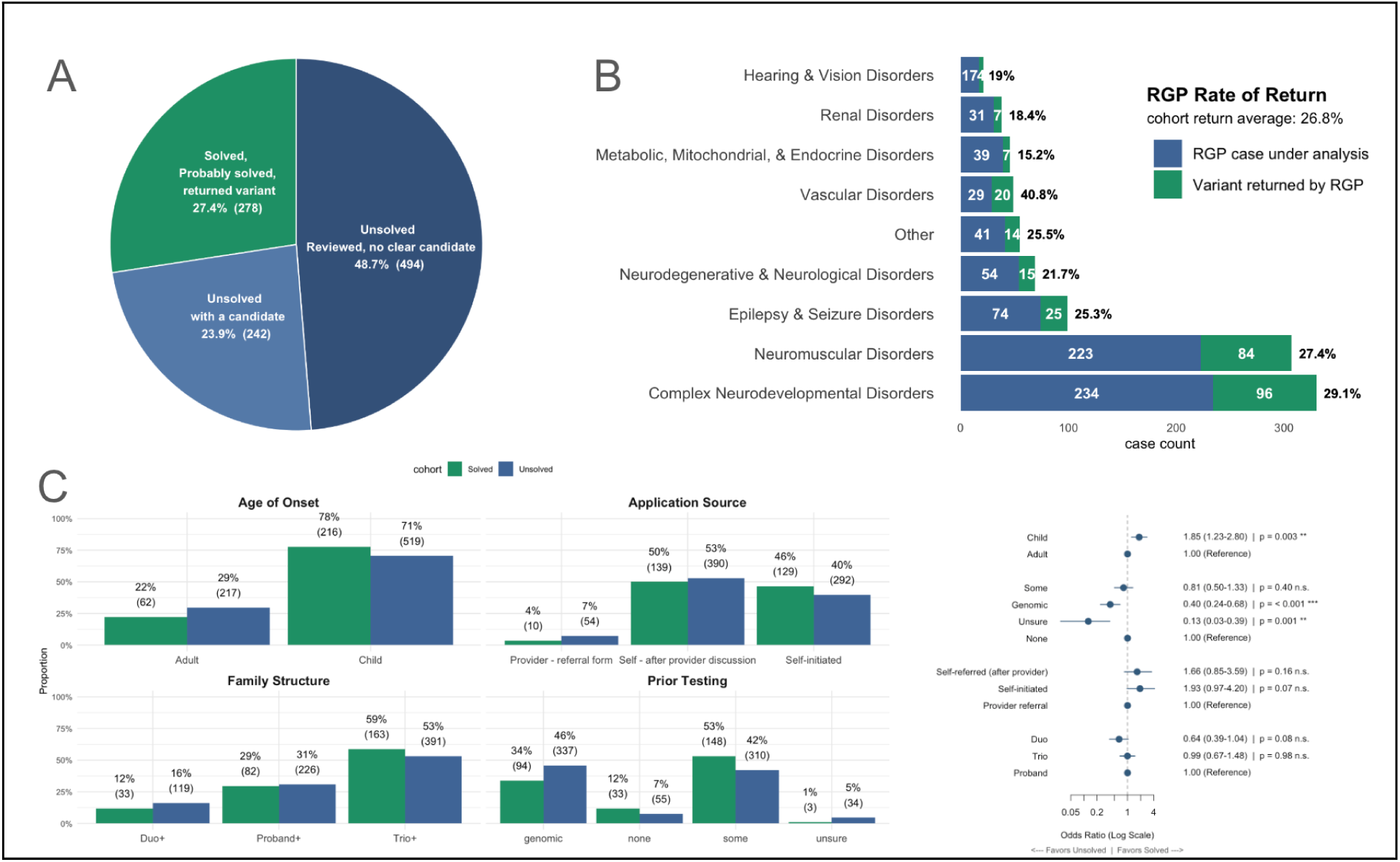
RGP cohort solved versus unsolved. (A) Pie chart of Solved, Unsolved, and Reviewed with candidate cases. (B) Breakdown of RGP family count by condition with the blue bar representing cases under analysis and the green bar representing cases with a primary returned result. The percentage of cases with a return per condition is shown. (C) Comparison of age of onset, application source, family structure and prior testing in solved versus unsolved cases. The odds ratio of each set is calculated with the reference group indicated.

Incidental findings unrelated to the proband’s reason for enrollment were identified and returned to five families, including four variants in ACMG SF v3.2 genes^36^ and one primary finding in a family member. Five VUS returns were ultimately re-curated and refuted as disease-causing based on subsequent biochemical testing (n=2), inconsistent familial segregation testing (n=2), or targeted clinical assessment (n=1). Families with only refuted or incidental findings (n=3) were subsequently re-grouped with “unsolved” or “unsolved with candidate” cases and remain in analysis.

Four families had primary returnable results in multiple genes: two probands with a possible dual diagnosis or blended phenotype (*ACADM*[MIM:201450]/heterozygous 12.3Mb 1p31.1p22.3 deletion and *CLTC* [MIM:617854]*/AGO1* [MIM:620292]), one family with two siblings with different causes of their neurodevelopmental phenotypes (*SCN2A* [MIM:613721] and *SATB2* [MIM:612313]), and one proband with a digenic *TTN/SRPK3* [MIM:301002] result^37^ (Table S2).

The RGP returnable result rate, excluding incidental and refuted findings, ranged from 15% to 40% depending on body system involvement (Figure 2B). While vascular conditions represented less than five percent of the total sequenced families, this category had the highest condition-specific return rate at 40.8% (20/49). This finding is likely due to RGP’s strategic partnership with the Cure HHT (Hereditary Hemorrhagic Telangiectasia) patient advocacy group, which included a disproportionate number of families with no prior genetic testing (33% versus 8% cohort average; Table S2). The returnable result rates for complex neurodevelopmental, neuromuscular, and other neurological phenotypes were next highest (Figure 2B). Intriguingly, sequenced family structure did not impact the likelihood of a returnable result nor were trio+ families more likely to have a candidate identified (Figure 2C and Figure S4). Factors significantly associated with receiving a returnable result through the study included age of disease onset and prior genetic testing (Figure 2C).

Undiagnosed families frequently pursued multiple avenues to end their diagnostic odyssey, including the pursuit of concurrent clinical testing or research opportunities. In line with this, 54 of the 272 families (20%) who received a primary returnable result through RGP obtained the same findings through parallel clinical testing (17%) or other research studies (3%). Additionally, six families obtained diagnoses clinically after enrollment involving variants that were not detectable through RGP GS analysis. Further investigations into the missed variants reaffirmed known limitations in the accurate calling of complex variants in short-read GS data, specifically difficulties in the calling of *FGF14*-Related Ataxia GAA repeat expansions [MIM:620174], *FSHD1* macrosatellite repeat contractions [MIM:158900], and mosaic deletions (in *CDKL5* [MIM:300203] and *PHACTR1* [MIM:608723]), although both mosaic deletions were detected through clinical GS. Similarly, three families were solved by outside biomedical researchers testing their latest analytical workflows and methodologies on our dataset; we facilitated these diagnoses by rapidly uploading genomic data to controlled access portals, such as NIH’s dbGaP/AnVIL and the Broad Institute’s DUOS, or by participating in the Critical Assessment of Genome Interpretation (CAGI) RGP challenge^38^. These findings involved complex interpretation and variant calling and included a diagnostic balanced translocation disrupting *ENG* [MIM:187300] (manuscript in preparation), intronic splice variants in *ASNS* [MIM:615574] and *TCF4* [MIM:610954]^38^, and a deletion in the 5’UTR of *AHDC1* [MIM:615829]^39^.

### Clinical Testing Gaps

Assessing the variants in returnable cases, 49% of families with a primary returnable result (134/272) could have had the result identified through clinical genetic testing with technologies and knowledge available at the time of enrollment. The majority of these results (110/134; 82%) met Solve/Probable Solve criteria without additional generated evidence or research technologies specific to this study. Research interventions as described below added supportive evidence for variant pathogenicity for the remaining 18% of cases (24/134).

The majority of these families (107/134) had a primary returnable result that would have reasonably been expected to be detected and reported had a comprehensive condition-specific panel or clinical ES been performed. The majority of these families (74/107) self-reported prior genetic testing that did not include the causal gene or necessary resolution to detect the causal variant. The remaining families (33/107) had no prior genetic testing performed, almost half of whom (15/33) carried a clinical diagnosis of a well-characterized monogenic condition including HHT, Hermansky-Pudlak syndrome, Sotos syndrome, Crouzon syndrome, or Gitelman syndrome.

The remaining families (27/134) had clinical ES or a clinical panel containing the relevant gene performed prior to enrollment, but the causative variant(s) were not reported. Clinically missed variant types were diverse and included mitochondrial variants, SNVs, indels, and SVs. Possible explanations for why the variants were not ascertained on prior testing were identified in 21 cases; structural variants, splice variants in the extended exon flanking region (3-20 bases from the intron/exon boundary), and variants relating to a gene discovery occurring 6-24 months before clinical testing accounted for the majority. (Figure 3). While the missed variant types are currently detectable by most clinical ES tests, exome capture kits, calling pipelines, and reportability criteria have varied over time and by lab. Of note, raw data from the clinical tests were not available for review to confirm they were in fact detected, versus missed due to technical limitations of the platforms or analytical pipelines.

**Figure 3:**
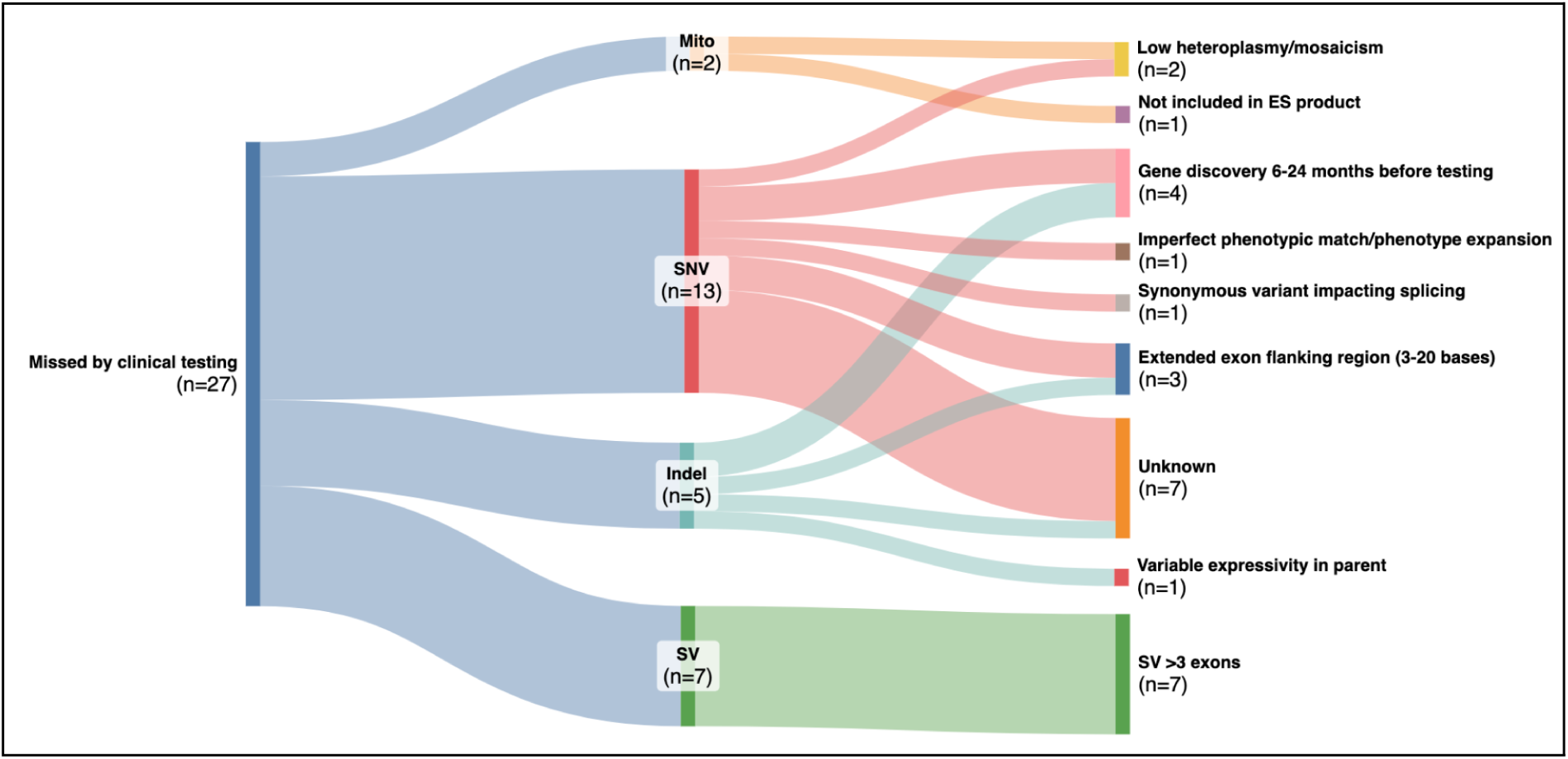
Variants missed by clinical panel and/or exome testing. Breakdown of variants missed by clinical testing by variant type with possible reasons why undetected or not reported. Extended exon flanking region variants occur in the intronic region 3-20 bases from the intron/exon boundary. Abbreviations: Mito, mitochondrial DNA variant; SNV, single nucleotide variant; Indel, insertion/deletion variant; SV, structural variant.

### Advancing Diagnosis through Research

The majority of primary returnable results required additional case evidence or data generation beyond ES (58.5%; 159/272). The additional data and methods utilized were classified into three non-exclusive categories: Interpretation, Technology, and Data Sharing (Table 2 & Figure 4).

**Figure 4:**
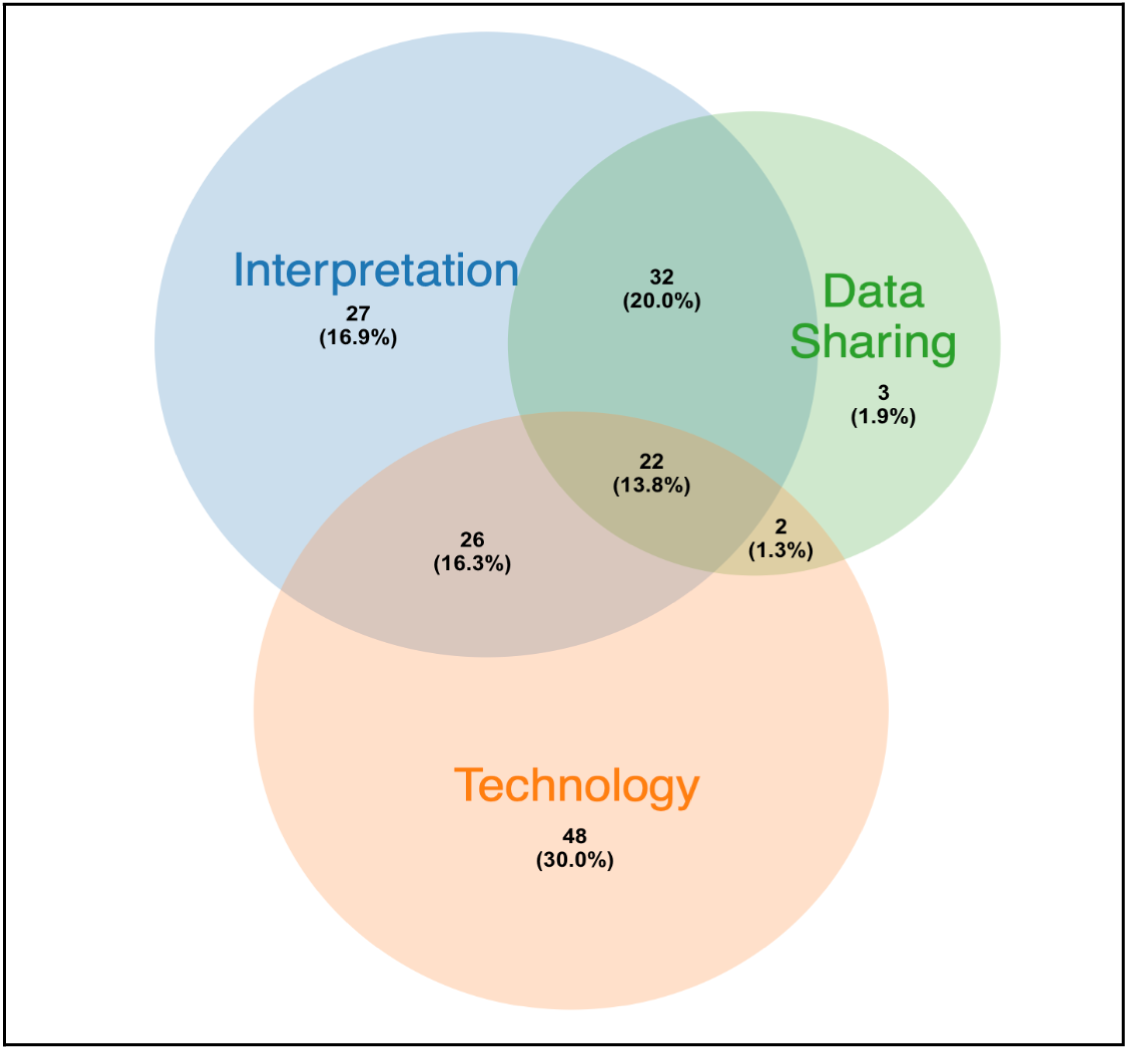
Research interventions. Venn diagram of research interventions leading to returnable results. The additional data and methods utilized are divided into three non-exclusive categories: Interpretation, Technology, and Data Sharing.

#### Interpretation - the gathering of data and case information to resolve uncertainty

Comprehensive curation of literature and review of medical records was needed to support variant or gene pathogenicity to increase confidence in variant return for 107 cases. Deep review of candidate genes for novel gene-disease associations or novel mechanisms account for 51% of cases involving any type of research intervention (81/159), highlighting the critical role of research in gene discovery and phenotype expansion. Resolution of VUSs in known genes was less frequent. The application of novel analytic tools assessing variant consequence contributed to the identification of 21 returnable variants, specifically deep intronic or cryptic splice variants identified with the help of newer *in silico* predictors such as SpliceAI^40^ or Pangolin^41^. In cases where specialized variant prioritization tools alone were not enough to resolve uncertainty, additional functional evidence was generated to support variant causality.

#### Technology - generation of new sequencing or functional data to support causality

Short-read GS and/or additional sequencing and functional data was required for 36% of primary returnable results. Similar to our prior report that included a partially overlapping cohort (39% of RGP cases reported here were included in Wojcik et al (2024)^30^, this study found a 6.8% increased yield with GS over ES (69 of 1014), including, but not limited to: the identification of intronic variants >20 bases from the intron/exon boundary; variants in non-coding genes, TREs, and complex SVs. This additional yield may be a slight underestimate, as some of the 27 variants missed by clinical testing may not have been detectable by exome/panel due to technical limitations and would thus have required GS. Additional sequencing technologies and functional data supporting variant pathogenicity was instrumental in 29% of cases (n=46) with RNA-seq and targeted high-throughput splicing or minigene assays for splice variants being the most fruitful. Segregation data beyond the biological parents, in both affected and unaffected family members, supported variant pathogenicity in 16% of all research intervention cases to contribute to segregation counts and confirm phase.

#### Data Sharing - leveraging variant- and gene-level matching and sharing de-identified data to advance diagnosis

Matchmaker Exchange via gene and variant level matching with the *seqr* platform^28^, and communication with external researchers with gene-specific knowledge and/or specialized analytic methods, were major factors in returnable results for 58 cases highlighting the importance of data sharing. In a smaller set of additional cases, primary returnable variants were first flagged by external users through controlled data access via the CAGI challenge^38^, Broad’s DUOS repository, or the GREGoR Consortium dataset on AnVIL. Open access ClinVar entries led to contact from researchers with additional case counts based on shared variants and clinical overlap. Our most prolific data sharing success was the discovery of ReNU syndrome and other disease-causing small nuclear RNAs (n=11, *RNU4-2* [MIM:620851] and *RNU2-2* [MIM:621304])^42–44^. Altogether, data sharing efforts led to RGP families being included in over 30 rare disease publications focused on novel gene discovery or deeper characterization of the phenotypic and variant spectrum of known genetic disorders (Table 3).

## DISCUSSION

### RGP advances the reach of genomic sequencing

The success of the Rare Genomes Project demonstrates that a direct-to-patient research model is critical to bridge the diagnostic gap for rare disease families. Our results also deepen the understanding of the genetic basis of many monogenic conditions. Strategies such as self-referral, fully remote participation, and partnerships with disease-specific patient advocacy groups and providers expanded the reach of our rare disease genomic research beyond that of traditional clinic-based models. The returnable result rate from 1,014 families, most of whom had prior clinical testing, reached 27%, with at least 6% of solved cases requiring GS technology, aligning with prior reports^30,45,46^. Outreach aimed at enrolling demographics currently underrepresented in clinic-based genomic studies across the U.S. evolved throughout the study duration and included techniques such as mobile phlebotomy and an expansion of the available languages on applications and during consent sessions. Our progress in equitable recruitment was previously documented and is the focus of ongoing research^47^.

### Research fills gaps in clinical reporting

Beyond serving as a “last resort” for undiagnosed families who have exhausted clinical options, we found genomic research programs play an important role as a primary access point to genetic testing for families not sufficiently served by the current healthcare infrastructure. While most sequenced families reported some measure of clinical genetic testing prior to enrollment (88%), the 45% percent with negative or inconclusive non-ES/GS results did not have follow-up genomic-level testing. The families with no prior genetic testing had the highest returnable results rate at 38.5% (versus 21.6% for those with prior ES/GS), and included 27 families with a diagnostic finding that could have been discovered via routine testing for a well-characterized monogenic condition. Forty-two percent of the families with no prior testing had access issues as evidenced by provider recommendation of RGP, lack of testing as discussed in clinic notes, or explicitly stated challenges with insurance coverage, with this being an underestimate as it was not a question asked of all applicants. Together these observations demonstrate that although coverage and access to genetic testing has improved, there is a clear need for continued progress in the education, implementation, and policy fronts of genomic medicine^7^.

We were able to uncover some addressable blind spots in clinical genetic testing. While over half of the solved families required sequencing techniques beyond ES or interpretation expertise, 44% could have been diagnosed at the time of enrollment with access to and optimized use of clinical ES. Suggested technical adjustments to ES analysis for increased diagnostic yield include the evaluation of intronic variants to +/- 20 nucleotides from the splice site along with the use of splice algorithms and variant prioritization tools, and the rapid implementation of updated gene-disease associations coupled with automated reanalysis pipelines to catch high-impact variants in newly characterized disease-genes. Additionally, all clinical ES tests should routinely interrogate exome data for copy number variants, mitochondrial DNA variants, and, for neuromuscular cases, the *SMN1/SMN2* locus using current best-in-class analysis tools and capture methods^48–50^.

### Research advances diagnostic potential through novel gene discovery

There are over 200 new disease-genes characterized every year, including many with clear diagnostic and medical management implications^10,51^. Although it is important for clinical labs offering ES to identify, share, and report variants in genes of unknown significance to increase the speed of novel gene discovery, this is not performed by many clinical laboratories ^52^. This study has demonstrated a clear increase in diagnostic potential through research-based novel disease-gene discovery. Thus far, the program has submitted 246 rare variants in candidate genes identified in undiagnosed individuals to platforms such as Matchmaker Exchange, and 34 RGP families have been featured in seminal publications defining new gene-disease relationships (Table 2), with many additional publications in progress. In total, 69 RGP returns involve variants in genes validated as a novel-disease gene, established through case collection and experimental evidence, after proband enrollment. This underscores the critical value of data sharing and systematic re-analysis, as well as highlights how engaging families in research helps delineate genes involved in disease and provide depth of disease spectra. Furthermore, it benefits the broader medical field as these newly established gene-disease associations are integrated into clinical genetic testing workflows globally thus impacting families beginning their diagnostic journey as well as families undergoing data re-analysis.

With increasing insurance coverage for GS, consideration can be given to making clinical GS a first-tier test for additional medical indications as has already been recommended for distinct patient populations^53–56^. For example, our experience with undiagnosed families with neuromuscular disease, a specialty where gene panels are typical^57^, we found a 27% solve rate that included diverse variant types, a subset of which required GS, and several discoveries involving phenocopies, phenotype expansions, variable expressivity. Expanding the first-tier test for this patient population to GS would result in not only reduced time until diagnosis but also negate the need for reflex testing which is often not covered. Panel test content and ES capture kit designs struggle to keep up with current gene-disease relationships, do not include deep intronic or non-coding regions implicated in disease, and, specifically for panels, do not have the potential to contribute to novel disease-gene discovery. Alternatively, the diagnostic gain for short-read GS has been proven across several independent studies, albeit modest at 6-9%, but more importantly holds the potential for non-coding interrogation and ongoing re-analysis, two areas shown to be especially fruitful for diagnosis^35,42–44,58^. The added yield for long-read sequencing over a best-in-class short-read GS remains limited^26,59,60^.

### Research remains an essential driver of innovation and knowledge

RGP demonstrates that research-based contributions in the areas of interpretation, technology, and/or data sharing substantively contributed to the majority of returnable results (58%), with half of these cases involving contribution from more than one area. Whereas clinical laboratories have targeted turn-around times for genetic tests and evaluate findings against existing gene-variant knowledge bases, the indefinite timeline of research allows for iterative re-analysis using a variety of novel tools and algorithms, targeted phenotyping, wet lab investigations and functional validation, and assessment of phenotype expansions and novel gene discoveries. Research programs like RGP have the bandwidth to solicit and share experimental findings through preprints, publications, and database submissions (e.g., ClinVar, GenCC) for ingestion into clinical pipelines, which leads to continual improvement to clinical diagnostic rates and medical knowledge. With thousands of disease-gene relationships yet to be elucidated^10^, novel gene-disease relationship discovery and phenotype/genotype expansions continue to remain core aims of rare disease research and necessary to advance genomic medicine.

Even in an ideal medical system where access to genomic sequencing was available for every patient, there would remain a critical need for genomic research programs to drive discovery, innovate on complementary methodologies, and obtain explicit consent for the broad use of human genomic data in the advancement of genomic health. The need for research institutions to accelerate the use of new techniques and methodologies and to give unbiased assessment on clinical utility is present now more than ever, especially in the age of competing and black-boxed artificial intelligence. Academic research is able to design pilot studies and quickly test the plethora of new techniques, analysis workflows, and AI predictors on well-studied datasets for clinical utility and scalability, as exemplified by the RGP CAGI experiments^38^. Clinical labs are less able and not incentivized to pivot or provide unbiased research with assessing new tools given low financial margins, consent restrictions, and rapid turn-around time requirements.

RGP housed at the Broad Institute of MIT and Harvard has established collaborations across global medical research programs and industry. This unique environment allowed for the application of a variety of sequencing and functional techniques to selected subsets of participants in order to assess the clinical utility of each technique for rare disease diagnosis. Our study found GS, RNA-seq, high-throughput splicing assays, and familial segregation testing to be the most powerful techniques to identify the causal genes and variants in individuals with undiagnosed genetic disease. Similarly, targeted use of tissue-appropriate RNA-seq, methylation episignature testing, and splicing validation assays for candidate variants of uncertain significance were powerful in resolving variant pathogenicity.

While our study highlights the critical role that disease-agnostic rare disease research programs continue to play in the discovery of novel causes of rare disease and the diagnosis of individuals with these diseases, funding for this work is at an all-time low with programs struggling to stay solvent. In the U.S., the Undiagnosed Diseases Network has struggled to remain active^61^, and the GREGoR consortium is shifting towards a focus on evaluating technologies and analysis methods on existing datasets with a limited number of new cases. New initiatives are needed to continue to drive novel gene-disease discovery and diagnosis rates for the many families who remain on their diagnostic odyssey.

### Challenges and limitations

This study has faced challenges related to being outside of a medical system, including difficulties sustaining steady recruitment, obtaining medical records, identifying discrepancies in self-reported phenotype and prior testing data, and coordinating appropriate result disclosure and counseling. Self-referral was shown to be a viable method of recruitment based on volume and had a slightly higher diagnostic yield than provider-referred families. Of note, assessment for the likelihood of a monogenic etiology was critical given the significant number of self-referred applicants who were deemed not eligible to participate.

We expected the ability to self-refer and participate remotely would render RGP accessible to any eligible family within the US. However, as previously described by our team, our subject population did not mirror US demographics. Targeted efforts did modestly improve the engagement of medically underserved populations in later years of the study, demonstrating a critical need for active, intentional outreach to underrepresented groups to ensure equitable access to the benefits of genomic research^47^. Improved capture of demographic characteristics will help identify gaps from which suitable interventions can be devised^62^. Recent efforts have been successful in intentionally recruiting a more diverse population in the All of Us Research Program but required extensive targeted efforts to achieve these goals^63^. Similarly targeted efforts focused on rare disease cohort recruitment are needed but will likely have more challenging barriers given existing disparities in subspecialty care observed in the US healthcare system^64–67^, a step often necessary to recognize rare disease symptoms with a likely genetic basis.

The majority of applications were from families who heard about RGP from other families, advocacy organizations or medical providers, suggesting that having a trusted source recommend the study was a factor in the decision to apply, which is particularly important for research-based studies conducted outside of a clinical setting. One effective recruitment strategy was partnership with rare disease patient advocacy organizations, such as those supported by the Rare As One Network. Our experience demonstrated that these groups are trusted resources for families and their existing networks of expertise help identify and reach the patients most likely to benefit from research opportunities as well as provide post-diagnosis support.

## Conclusion

The Rare Genomes Project (RGP) demonstrates that research-based genomic programs are not merely a supplement to clinical medicine, but are an essential driver of innovation in genomic diagnostics^28,35^. In addition to closing gaps in clinical test accessibility, research projects like RGP apply the latest in genomic technology, interpretation, and data sharing to drive discovery, evaluate diagnostic approaches, and inform best practices with the ultimate goal of shortening the diagnostic odyssey for families living with and affected by rare diseases.

Over its initial 9 years, RGP provided answers for 272 undiagnosed families and contributed to the discovery of at least 35 new gene-disease associations. The study created a rare disease dataset that continues to empower rare disease research for ongoing gene discovery and technological advancements. Data and samples supplied by solved and unsolved participants^34^ will fuel future innovations in gene discovery and rare disease research, generating the essential data needed to advance clinical pipelines for the improvement of genomic diagnosis.

## Data Availability

All data produced are available online by controlled access through AnVIL as part of the GREGoR (Genomics Research to Elucidate the Genetics of Rare diseases) Consortium (phs003047); the Broad Institute Data Use Oversight System under dataset name (RGP_Rehm_RareDisease_WGS - ID DUOS_000008; and through the GREGoR variant browser. Variant classifications have been submitted to ClinVar.

https://www.ncbi.nlm.nih.gov/projects/gap/cgi-bin/study.cgi?study_id=phs003047.v5.p4

https://duos.broadinstitute.org/?redirectTo=%2Fdatalibrary

https://www.ncbi.nlm.nih.gov/clinvar/submitters/506627/

https://variants.gregorconsortium.org/

## ACKNOWLEDGMENTS

Recruitment, sequencing, analysis, and return of results were provided by the Broad Institute of MIT and Harvard Center for Mendelian Genomics (Broad CMG) and were funded by the National Human Genome Research Institute (NHGRI) grants UM1HG008900 (CMG program with additional support from the National Eye Institute, and the National Heart, Lung and Blood Institute), U01HG0011755 (GREGoR consortium), R21HG012397, R01HG009141, and R01HG013986, and support from the National Institute of Diabetes and Digestive and Kidney Diseases grant K23DK114551, and in part by Illumina and the Chan Zuckerberg Initiative Donor-Advised Fund at the Silicon Valley Community Foundation (funder DOI 10.13039/100014989) grants 2019-199278, 2020-224274, 2022-316726 (https://doi.org/10.37921/236582yuakxy). Additional study funding was provided by the Muscular Dystrophy Association (MDA), Illumina, Inc., Pacific Biosciences of California, Inc., Shriners Hospitals for Children, the Thrasher Research Fund, and the Mathers Foundation. An estimated 85% of this work was funded by federal funds and 15% funded by the other listed sources. The content is solely the responsibility of the authors and does not necessarily represent the official views of the funding agencies.

Our deepest thanks to the participating families, our dedicated advocacy partners, the Rare As One Network, and the Broad Institute’s Broad Clinical Labs/Genomics Platform, Data Sciences Platform, and Data Donation Platform.

## AUTHOR CONTRIBUTIONS

AODL, HLR, DGM, MET, and HB conceptualized the study. AODL, HLR, DGM, JHL, MCO, HB, KES, and MET developed the study plan. BB, JG, KL, HS, BW, and MHW developed and implemented software and data algorithms. MA, CAT, KMB, CMC, SC, LE C, SD, EME, VSG, JG, EG, GL, AKL, JM, CM, EO, BO, IAOO, LP, SP, LR, ASJ, RS, ES, KMS, SLS, BW, RW, and MHW conducted data analysis. SA, KMB, SC, SD, EG, NL, AKL, BEM, EM, AN, AODL, EO, MCO, IAOO, LP, HLR, LR, RS, ES, JS, GS, MS, GEV, BW, and RW conducted study investigations. SA, NL, BEM, EM, AN, MCO, JS, GS, CEW, and MHW compiled study resources or performed participant recruitment. CAT, SMB, CG, KLarichhia, GL, SJ, KR, MSB, KMS, and MHW performed data curation. SD, AODL, MCO, and HLR wrote the original draft. CM, AN, LR, ES, KMS, MET, MU, and GEV provided manuscript review and edits. SD, MCO, and AP provided data visualization. CAT, SMB, HB, SG DGM, AODL, MCO, HLR, MET, and GEV supervised study activities. SA, BEM, AN, AODL, MCO, AP, HLR, MU, and RW coordinated study activities/performed project management. DGM, AODL, HLR, MET, and MU acquired financial support.

## DATA CODE AND AVAILABILITY

Controlled access to sequence and phenotype data is available through NHGRI’s data-sharing platform AnVIL (Analysis, Visualization, and Informatics Lab-space) as part of the GREGoR (Genomics Research to Elucidate the Genetics of Rare diseases) Consortium (phs003047), as well as through the Broad Institute’s Data Use Oversight System (DUOS; https://duos.broadinstitute.org/) under dataset name ‘RGP_Rehm_RareDisease_WGS’ (ID DUOS_000008). Variants in RGP can be queried through Variant-Level Matching (VLM) through the *seqr* platform (seqr.broadinstitute.org), part of the federated VLM platform organized through the Global Alliance for Genomics and Health^68^ as well as through the GREGoR variant browser (https://variants.gregorconsortium.org/). Steps for full data access can be found on the respective AnVIL, DUOS, and GREGoR websites.

## REFERENCES

1. Tifft, C.J., and Adams, D.R. (2014). The National Institutes of Health undiagnosed diseases program. Curr. Opin. Pediatr. 26, 626–633.

2. 100,000 Genomes Project Pilot Investigators, Smedley, D., Smith, K.R., Martin, A., Thomas, E.A., McDonagh, E.M., Cipriani, V., Ellingford, J.M., Arno, G., Tucci, A., et al. (2021). 100,000 genomes pilot on rare-disease diagnosis in health care - preliminary report. N. Engl. J. Med. 385, 1868–1880.

3. Wojcik, M.H., Reuter, C.M., Marwaha, S., Mahmoud, M., Duyzend, M.H., Barseghyan, H., Yuan, B., Boone, P.M., Groopman, E.E., Délot, E.C., et al. (2023). Beyond the exome: What’s next in diagnostic testing for Mendelian conditions. Am. J. Hum. Genet. 110, 1229–1248.

4. Bauskis, A., Strange, C., Molster, C., and Fisher, C. (2022). The diagnostic odyssey: insights from parents of children living with an undiagnosed condition. Orphanet J. Rare Dis. 17, 233.

5. Splinter, K., Adams, D.R., Bacino, C.A., Bellen, H.J., Bernstein, J.A., Cheatle-Jarvela, A.M., Eng, C.M., Esteves, C., Gahl, W.A., Hamid, R., et al. (2018). Effect of Genetic Diagnosis on Patients with Previously Undiagnosed Disease. N. Engl. J. Med. 379, 2131–2139.

6. Fraiman, Y.S., and Wojcik, M.H. (2021). The influence of social determinants of health on the genetic diagnostic odyssey: who remains undiagnosed, why, and to what effect? Pediatr. Res. 89, 295–300.

7. Jobanputra, V., Schroeder, B., Rehm, H.L., Shen, W., Spiteri, E., Nakouzi, G., Taylor, S., Marshall, C.R., Meng, L., Kingsmore, S.F., et al. (2024). Advancing access to genome sequencing for rare genetic disorders: recent progress and call to action. NPJ Genom. Med. 9, 23.

8. Rehm, H.L., Alaimo, J.T., Aradhya, S., Bayrak-Toydemir, P., Best, H., Brandon, R., Buchan, J.G., Chao, E.C., Chen, E., Clifford, J., et al. (2023). The landscape of reported VUS in multi-gene panel and genomic testing: Time for a change. Genet. Med. 25, 100947.

9. Fowler, D.M., and Rehm, H.L. (2024). Will variants of uncertain significance still exist in 2030? Am. J. Hum. Genet. 111, 5–10.

10. Bamshad, M.J., Nickerson, D.A., and Chong, J.X. (2019). Mendelian Gene Discovery: Fast and Furious with No End in Sight. Am. J. Hum. Genet. 105, 448–455.

11. Gahl, W.A., Mulvihill, J.J., Toro, C., Markello, T.C., Wise, A.L., Ramoni, R.B., Adams, D.R., Tifft, C.J., and UDN (2016). The NIH Undiagnosed Diseases Program and Network: Applications to modern medicine. Mol. Genet. Metab. 117, 393–400.

12. Baxter, S.M., Posey, J.E., Lake, N.J., Sobreira, N., Chong, J.X., Buyske, S., Blue, E.E., Chadwick, L.H., Coban-Akdemir, Z.H., Doheny, K.F., et al. (2022). Centers for Mendelian Genomics: A decade of facilitating gene discovery. Genet. Med. 24, 784–797.

13. Dawood, M., Heavner, B., Wheeler, M.M., Ungar, R.A., LoTempio, J., Wiel, L., Berger, S., Bernstein, J.A., Chong, J.X., Délot, E.C., et al. (2025). GREGoR: accelerating genomics for rare diseases. Nature 647, 331–342.

14. Cloney, T., Gallacher, L., Pais, L.S., Tan, N.B., Yeung, A., Stark, Z., Brown, N.J., McGillivray, G., Delatycki, M.B., de Silva, M.G., et al. (2022). Lessons learnt from multifaceted diagnostic approaches to the first 150 families in Victoria’s Undiagnosed Diseases Program. J. Med. Genet. 59, 748–758.

15. Bakur, K., Hamid, H., Alhaddad, B., Alfadhel, M., Alhashem, A., Eyaid, W., Alanzi, T., Al Mutairi, F., Alswaid, A., Ababneh, F., et al. (2025). Adult genomic medicine: lessons from a multisite study of 2700 patients. Genome Med. 17, 105.

16. Boycott, K.M., Hartley, T., Kernohan, K.D., Dyment, D.A., Howley, H., Innes, A.M., Bernier, F.P., Brudno, M., and Care4Rare Canada Consortium (2022). Care4Rare Canada: Outcomes from a decade of network science for rare disease gene discovery. Am. J. Hum. Genet. 109, 1947–1959.

17. Brazilian Rare Genomes Project Consortium. Electronic address, and Brazilian Rare Genomes Project Consortium (2026). Genome sequencing for the diagnosis of rare disorders: The Brazilian Rare Genomes Project. HGG Adv. 7, 100624.

18. Wright, C.F., Fitzgerald, T.W., Jones, W.D., Clayton, S., McRae, J.F., van Kogelenberg, M., King, D.A., Ambridge, K., Barrett, D.M., Bayzetinova, T., et al. (2015). Genetic diagnosis of developmental disorders in the DDD study: a scalable analysis of genome-wide research data. Lancet 385, 1305–1314.

19. Curic, E., Ewans, L., Pysar, R., Taylan, F., Botto, L.D., Nordgren, A., Gahl, W., and Palmer, E.E. (2023). International Undiagnosed Diseases Programs (UDPs): components and outcomes. Orphanet J. Rare Dis. 18, 348.

20. Caetano-Anollés, D. (2026). Discovering, genotyping, and annotating structural variants with the Genome Analysis Toolkit (GATK). Methods Mol. Biol. 2981, 107–117.

21. Basu, S., Xie, X., Uhler, J.P., Hedberg-Oldfors, C., Milenkovic, D., Baris, O.R., Kimoloi, S., Matic, S., Stewart, J.B., Larsson, N.-G., et al. (2020). Accurate mapping of mitochondrial DNA deletions and duplications using deep sequencing. PLoS Genet. 16, e1009242.

22. Dolzhenko, E., Deshpande, V., Schlesinger, F., Krusche, P., Petrovski, R., Chen, S., Emig-Agius, D., Gross, A., Narzisi, G., Bowman, B., et al. (2019). ExpansionHunter: a sequence-graph-based tool to analyze variation in short tandem repeat regions. Bioinformatics 35, 4754–4756.

23. Weisburd, B., Sharma, R., Pata, V., Reimand, T., Ganesh, V.S., Austin-Tse, C., Osei-Owusu, I., O’Heir, E., O’Leary, M., Pais, L., et al. (2024). Detecting missed diagnoses of spinal muscular atrophy in genome, exome, and panel sequencing datasets. medRxiv. 10.1101/2024.02.11.24302646.

24. Awamleh, Z., Goodman, S., Kallurkar, P., Wu, W., Lu, K., Choufani, S., Turinsky, A.L., and Weksberg, R. (2022). Generation of DNA methylation signatures and classification of variants in rare neurodevelopmental disorders using EpigenCentral. Curr. Protoc. 2, e597.

25. Negi, S., Stenton, S.L., Berger, S.I., Canigiula, P., McNulty, B., Violich, I., Gardner, J., Hillaker, T., O’Rourke, S.M., O’Leary, M.C., et al. (2025). Advancing long-read nanopore genome assembly and accurate variant calling for rare disease detection. Am. J. Hum. Genet. 112, 428–449.

26. Sanchis-Juan, A., Mostovoy, Y., Stenton, S.L., Ganesh, V.S., Weisburd, B., Yenkin, A., Kurtas, N.E., Zhao, X., Shin, E., Boone, P.M., et al. (2026). Structural variant discovery and diagnostic impact in rare diseases from short-read and long-read sequencing. medRxiv. 10.64898/2026.06.22.26356238.

27. Brownstein, C.A., van der Made, C.I., Cabral, K., Rockowitz, S., Kang, D., Schieck, M., Pang, A.W.C., Robinson, J.M., Hastie, A.R., Chaubey, A., et al. (2025). Rare structural variants uncovered by optical genome mapping in multisystem inflammatory syndrome in children (MIS-C). Adv. Genet. (Hoboken) 6, e00023.

28. Pais, L.S., Snow, H., Weisburd, B., Zhang, S., Baxter, S.M., DiTroia, S., O’Heir, E., England, E., Chao, K.R., Lemire, G., et al. (2022). seqr: A web-based analysis and collaboration tool for rare disease genomics. Hum. Mutat. 43, 698–707.

29. Boycott, K.M., Azzariti, D.R., Hamosh, A., and Rehm, H.L. (2022). Seven years since the launch of the Matchmaker Exchange: The evolution of genomic matchmaking. Hum. Mutat. 43, 659–667.

30. Wojcik, M.H., Lemire, G., Berger, E., Zaki, M.S., Wissmann, M., Win, W., White, S.M., Weisburd, B., Wieczorek, D., Waddell, L.B., et al. (2024). Genome sequencing for diagnosing rare diseases. N. Engl. J. Med. 390, 1985–1997.

31. Riggs, E.R., Andersen, E.F., Cherry, A.M., Kantarci, S., Kearney, H., Patel, A., Raca, G., Ritter, D.I., South, S.T., Thorland, E.C., et al. (2020). Technical standards for the interpretation and reporting of constitutional copy-number variants: a joint consensus recommendation of the American College of Medical Genetics and Genomics (ACMG) and the Clinical Genome Resource (ClinGen). Genet. Med. 22, 245–257.

32. Richards, S., Aziz, N., Bale, S., Bick, D., Das, S., Gastier-Foster, J., Grody, W.W., Hegde, M., Lyon, E., Spector, E., et al. (2015). Standards and guidelines for the interpretation of sequence variants: a joint consensus recommendation of the American College of Medical Genetics and Genomics and the Association for Molecular Pathology. Genet. Med. 17, 405–424.

33. Strande, N.T., Riggs, E.R., Buchanan, A.H., Ceyhan-Birsoy, O., DiStefano, M., Dwight, S.S., Goldstein, J., Ghosh, R., Seifert, B.A., Sneddon, T.P., et al. (2017). Evaluating the Clinical Validity of Gene-Disease Associations: An Evidence-Based Framework Developed by the Clinical Genome Resource. Am. J. Hum. Genet. 100, 895–906.

34. Heavner, B.D., Wheeler, M.M., Bengtsson, J.D., Carvalho, C.M., Cheung, W.A., Conomos, M.P., Délot, E.C., DiTroia, S., Ganesh, V.S., Gogarten, S.M., et al. (2026). Building an interoperable rare disease multi-omic resource: The GREGoR Data Model and dataset. bioRxivorg. 10.64898/2026.05.15.725546.

35. Welland, M.J., Ahlquist, K.D., De Fazio, P., Austin-Tse, C., Pais, L., Wedd, L., Bryen, S., Rius, R., Franklin, M., Morrison, C., et al. (2026). Automated reanalysis of genomic data for rare disease diagnostics at scale. Nat. Med. 10.1038/s41591-026-04477-5.

36. Miller, D.T., Lee, K., Abul-Husn, N.S., Amendola, L.M., Brothers, K., Chung, W.K., Gollob, M.H., Gordon, A.S., Harrison, S.M., Hershberger, R.E., et al. (2023). ACMG SF v3.2 list for reporting of secondary findings in clinical exome and genome sequencing: A policy statement of the American College of Medical Genetics and Genomics (ACMG). Genet. Med. 25, 100866.

37. Töpf, A., Cox, D., Zaharieva, I.T., Di Leo, V., Sarparanta, J., Jonson, P.H., Sealy, I.M., Smolnikov, A., White, R.J., Vihola, A., et al. (2024). Digenic inheritance involving a muscle-specific protein kinase and the giant titin protein causes a skeletal muscle myopathy. Nat. Genet. 56, 395–407.

38. Stenton, S.L., O’Leary, M.C., Lemire, G., VanNoy, G.E., DiTroia, S., Ganesh, V.S., Groopman, E., O’Heir, E., Mangilog, B., Osei-Owusu, I., et al. (2024). Critical assessment of variant prioritization methods for rare disease diagnosis within the rare genomes project. Hum. Genomics 18, 44.

39. Bertrand, M., Shah, G., Pedersen, B.S., Schulz, A., Weise, A., Liehr, T., Huppke, P., DiTroia, S., Quinlan, A.R., Haack, T.B., et al. (2024). De novo AHDC1 deletions identified by genome sequencing in two individuals with Xia-Gibbs syndrome. Mol. Syndromol. 15, 389–397.

40. Jaganathan, K., Kyriazopoulou Panagiotopoulou, S., McRae, J.F., Darbandi, S.F., Knowles, D., Li, Y.I., Kosmicki, J.A., Arbelaez, J., Cui, W., Schwartz, G.B., et al. (2019). Predicting splicing from primary sequence with deep learning. Cell 176, 535–548.e24.

41. Zeng, T., and Li, Y.I. (2022). Predicting RNA splicing from DNA sequence using Pangolin. Genome Biol. 23, 103.

42. Leitão, E., Santini, A., Cogne, B., Essid, M., Athanasiadou, M., LaFlamme, C.W., Marijon, P., Bernard, V., Jousselin, K., Chatron, N., et al. (2026). Systematic analysis of snRNA genes reveals frequent RNU2-2 variants in dominant and recessive developmental and epileptic encephalopathies. Nat. Genet. 58, 782–797.

43. Rius, R., Blakes, A.J.M., Chen, Y., De Jonghe, J., Lecoquierre, F., Dawes, R., Cogne, B., Kim, H.C., Alvi, J.R., Amblard, F., et al. (2026). Biallelic variants in the noncoding RNA gene RNU4-2 cause a recessive neurodevelopmental syndrome with distinct white matter changes. Nat. Genet. 58, 761–773.

44. Chen, Y., Dawes, R., Kim, H.C., Ljungdahl, A., Stenton, S.L., Walker, S., Lord, J., Lemire, G., Martin-Geary, A.C., Ganesh, V.S., et al. (2024). De novo variants in the RNU4-2 snRNA cause a frequent neurodevelopmental syndrome. Nature 632, 832–840.

45. Alfares, A., Aloraini, T., Subaie, L.A., Alissa, A., Qudsi, A.A., Alahmad, A., Mutairi, F.A., Alswaid, A., Alothaim, A., Eyaid, W., et al. (2018). Whole-genome sequencing offers additional but limited clinical utility compared with reanalysis of whole-exome sequencing. Genet. Med. 20, 1328–1333.

46. Albuquerque, A.L.B., Dos Santos, G.G., Sadok, S.H., Antonello, B.B., de Jesus, L.M., de Carvalho, M.E.A., Mutarelli, A., and Ribeiro, P.V.Z. (2025). Diagnostic yield of genome sequencing versus exome sequencing in pediatric patients with rare phenotypes: A systematic review and meta-analysis. Am. J. Med. Genet. A 197, e64146.

47. Martinez, E., Serrano, J., Abouhala, S., Neale, A., VanNoy, G., Rehm, H.L., O’Leary, M., O’Donnell-Luria, A., and Wojcik, M.H. (2026). Equity-focused implementation to enhance access to rare disease genomic research and understand diverse perspectives. Genet. Med. 28, 101667.

48. Lemire, G., Sanchis-Juan, A., Russell, K., Baxter, S., Chao, K.R., Singer-Berk, M., Groopman, E., Wong, I., England, E., Goodrich, J., et al. (2024). Exome copy number variant detection, analysis, and classification in a large cohort of families with undiagnosed rare genetic disease. Am. J. Hum. Genet. 111, 863–876.

49. Weisburd, B., Sharma, R., Pata, V., Reimand, T., Ganesh, V.S., Austin-Tse, C., Osei-Owusu, I., O’Heir, E., O’Leary, M., Pais, L., et al. (2025). Diagnosing missed cases of spinal muscular atrophy in genome, exome, and panel sequencing data sets. Genet. Med. 27, 101336.

50. Stenton, S.L., Laricchia, K., Lake, N.J., Chaluvadi, S., Ganesh, V., DiTroia, S., Osei-Owusu, I., Pais, L., O’Heir, E., Austin-Tse, C., et al. (2025). Mitochondrial DNA variant detection in over 6,500 rare disease families by the systematic analysis of exome and genome sequencing data resolves undiagnosed cases. HGG Adv. 6, 100441.

51. Seaby, E.G., Rehm, H.L., and O’Donnell-Luria, A. (2021). Strategies to uplift novel Mendelian gene discovery for improved clinical outcomes. Front. Genet. 12, 674295.

52. Chong, J.X., Berger, S.I., Baxter, S., Smith, E., Xiao, C., Calame, D.G., Hawley, M.H., Rivera-Munoz, E.A., DiTroia, S., Genomics Research to Elucidate the Genetics of Rare Diseases (GREGoR) Consortium, et al. (2024). Considerations for reporting variants in novel candidate genes identified during clinical genomic testing. Genet. Med. 26, 101199.

53. Rodan, L.H., Stoler, J., Chen, E., Geleske, T., and Council on Genetics (2025). Genetic evaluation of the child with intellectual disability or global developmental delay: Clinical report. Pediatrics 156, e2025072219.

54. Manickam, K., McClain, M.R., Demmer, L.A., Biswas, S., Kearney, H.M., Malinowski, J., Massingham, L.J., Miller, D., Yu, T.W., Hisama, F.M., et al. (2021). Exome and genome sequencing for pediatric patients with congenital anomalies or intellectual disability: an evidence-based clinical guideline of the American College of Medical Genetics and Genomics (ACMG). Genet. Med. 23, 2029–2037.

55. Kulsirichawaroj, P., Chanvanichtrakool, M., Wattanadilokchatkun, P., Pho-Iam, T., Limwongse, C., Likasitwattanakul, S., Boonyapisit, K., Sanmaneechai, O., Nishino, I., Shotelersuk, V., et al. (2025). Next-generation sequencing for pediatric-onset neuromuscular disorders unresolved by conventional diagnostic methods. Pediatr. Res. 98, 2195–2202.

56. Shickh, S., Gutierrez Salazar, M., Zakoor, K.-R., Lázaro, C., Gu, J., Goltz, J., Kleinman, D., Noor, A., Khalouei, S., Mighton, C., et al. (2021). Exome and genome sequencing in adults with undiagnosed disease: a prospective cohort study. J. Med. Genet. 58, 275–283.

57. Ng, K.W.P., Chin, H.-L., Chin, A.X.Y., and Goh, D.L.-M. (2022). Using gene panels in the diagnosis of neuromuscular disorders: A mini-review. Front. Neurol. 13, 997551.

58. Ganesh, V.S., Riquin, K., Chatron, N., Yoon, E., Lamar, K.-M., Aziz, M.C., Monin, P., O’Leary, M.C., Goodrich, J.K., Garimella, K.V., et al. (2024). Neurodevelopmental disorder caused by deletion of CHASERR, a lncRNA gene. N. Engl. J. Med. 391, 1511–1518.

59. Hiatt, S.M., Lawlor, J.M.J., Handley, L.H., Latner, D.R., Bonnstetter, Z.T., Finnila, C.R., Thompson, M.L., Boston, L.B., Williams, M., Rodriguez Nunez, I., et al. (2024). Long-read genome sequencing and variant reanalysis increase diagnostic yield in neurodevelopmental disorders. Genome Res. 34, 1747–1762.

60. Ek, M., Kvarnung, M., Ten Berk de Boer, E., La Fleur, L., Ljöstad, L., Lyander, A., Faergeman, S.L., Drue, S.O., Thonberg, H., Nordgren, A., et al. (2026). Long-read genome sequencing enhances diagnostics of pediatric neurological disorders. Genome Med. 18, 12.

61. Kaiser, J. (2022). Funds dwindle for NIH program for puzzling cases. Science 377, 15.

62. Sinan, I., Johnston, M., and Marwaha, A. (2026). Best practices in demographic data collection for equity, diversity, and inclusion in rare disease research: A systematic review. Genet. Med., 102592.

63. Bianchi, D.W., Brennan, P.F., Chiang, M.F., Criswell, L.A., D’Souza, R.N., Gibbons, G.H., Gilman, J.K., Gordon, J.A., Green, E.D., Gregurick, S., et al. (2024). The All of Us Research Program is an opportunity to enhance the diversity of US biomedical research. Nat. Med. 30, 330–333.

64. Wojcik, M.H., Bresnahan, M., Del Rosario, M.C., Ojeda, M.M., Kritzer, A., and Fraiman, Y.S. (2023). Rare diseases, common barriers: disparities in pediatric clinical genetics outcomes. Pediatr. Res. 93, 110–117.

65. Halley, M.C., Halverson, C.M.E., Tabor, H.K., and Goldenberg, A.J. (2023). Rare disease, advocacy and justice: Intersecting disparities in research and clinical care. Am. J. Bioeth. 23, 17–26.

66. Serrano, J.G., O’Leary, M., VanNoy, G.E., Mangilog, B.E., Holm, I.A., Fraiman, Y.S., Rehm, H.L., O’Donnell-Luria, A., and Wojcik, M.H. (2023). Advancing understanding of inequities in rare disease genomics. Clin. Ther. 45, 745–753.

67. Tang, Z., Mis, E.K., and Lakhani, S.A. (2026). Referral route: a determinant of inequity for children with undiagnosed genetic diseases? Front. Genet. 17, 1692489.

68. Rodrigues, E. da S., Griffith, S., Martin, R., Antonescu, C., Posey, J.E., Coban-Akdemir, Z., Jhangiani, S.N., Doheny, K.F., Lupski, J.R., Valle, D., et al. (2022). Variant-level matching for diagnosis and discovery: Challenges and opportunities. Hum. Mutat. 43, 782–790.

69. Department of Agriculture, Economic Research Service. 2020 Rural-Urban Commuting Area Codes (2025).

